# Development of web-based quality-assurance tool for radiotherapy target delineation for head and neck cancer: quality evaluation of nasopharyngeal carcinoma

**DOI:** 10.1101/2021.02.24.21252123

**Authors:** Jun Won Kim, Joseph Marsilla, Michal Kazmierski, Denis Tkachuk, Shao Hui Huang, Wei Xu, John Cho, Jolie Ringash, Scott Bratman, Benjamin Haibe-Kains, Andrew Hope

## Abstract

**Purpose:** We developed *QUANNOTATE*, a new web-application for rapid review of radiotherapy (RT) target volumes, and used it to evaluate the relationship between target delineation compliance with the international guidelines and treatment outcomes in nasopharyngeal carcinoma (NPC) patients undergoing definitive RT.

**Methods and Materials:** The dataset used for this study consists of anonymized CT simulation scans, RT structures, and clinical data of 332 pathologically confirmed NPC patients treated with intensity-modulated RT between July 2005 and August 2017. We imported the contours of intermediate risk clinical target volumes of the primary tumor (IR-CTVp) receiving 56 Gy into *QUANNOTATE*. We determined inclusion of anatomic sites within IR-CTVp in accordance with 2018 International guideline for CTV delineation for NPC and correlated the results with time to local failure (TTLF) using Cox-regression.

**Results:** At a median follow-up of 5.6 years, 5-year TTLF and overall survival rates were 93.1% and 85.9% respectively. The most frequently non-guideline compliant anatomic sites were sphenoid sinus (n = 69, 20.8%), followed by cavernous sinus (n = 38, 19.3%), left and right petrous apices (n = 37 and 32, 11.1% and 9.6%), clivus (n = 14, 4.2%), and right and left foramen rotundum (n = 14 and 12, 4.2% and 3.6%). Among 23 patients with a local failure (6.9%), the number of non-compliant cases were 8 for sphenoid sinus, 7 cavernous sinus, 4 left and 3 right petrous apices, and 2 clivus. Compared to conforming cases, cases which did not contour the cavernous sinus had a higher local failure (LF) rate (89.1% vs 93.6%, p= 0.013). Multivariable analysis confirmed that lack of cavernous sinus contouring was prognostic for LF.

**Conclusions:** *QUANNOTATE* allowed rapid review of target volumes in a large patient cohort. Despite an overall high compliance with the international guidelines, undercoverage of the cavernous sinus was correlated with LF.

## Introduction

Intensity-modulated radiotherapy (IMRT) is a current standard option for the treatment of head and neck cancer (HNC) and often allows effective sparing of organs at risk (OARs). Dose-painting via IMRT allows delivery of non-uniform doses to clinical target volumes (CTVs) of different recurrence risks, thus improving therapeutic ratio.^1^ In the phase II Radiation Therapy Oncology Group (RTOG) 0225 trial of IMRT with or without chemotherapy for nasopharyngeal carcinoma (NPC), CTV70 or high risk CTV (HR-CTV) was defined as gross tumor volume (GTV) + 5-mm margin, and CTV59.4 or intermediate risk CTV (IR-CTV) as CTV70 + 5-mm margin plus areas at risk for microscopic involvement. The recommended coverage for microscopic involvement included the entire nasopharynx, retropharyngeal nodal regions, skull base, clivus, pterygoid fossae, parapharyngeal space, sphenoid sinus, the posterior third of the nasal cavity/maxillary sinuses and the pterygopalatine fossae.^2^ The extent of IR-CTV in NPC treatment is related to marginal local recurrences^3^ and development of radiation toxicity including xerostomia and swallowing difficulty,^4^ yet inter-observer variation in delineating IR-CTV can be substantial.^5^ With advancement of conformal radiotherapy (RT) techniques and reduction of target margins, adequate target coverage has become a critical issue, and consensus guidelines for delineation of neck node levels^6^ and CTVs for conformal RT for nasopharynx and other head and neck tumor sites have been published.^7,8^

RT quality assurance (QA) is particularly important for HNC treatment, especially in the conformal RT era, due to the complexity of RT target volumes and multiple adjacent OARs.^9^ In a multicenter, international phase III randomized trial, Peters et al noticed that, “Centers treating only a few patients are the major source of quality problems.”^10^ Clinical studies rely on assumption and/or maintenance of adequate QA; however, limitations in time and human resources to review large patient cohorts often result in compliance issues. Multi-institutional prospective RT trials require central review of RT target delineation, yet only the first few cases are submitted and reviewed for participation in many trials.^11^ Patterns-of-radiotherapy-practice studies often rely on survey questionnaires,^12^ instead of visually inspecting the RT structures, target volumes and OARs.

To address these issues, we developed *QUANNOTATE*, a new quality assurance tool that allows rapid review of large numbers of RT target volumes in an easily accessible format without requiring access to the RT planning system (https://www.quannotate.com). Using *QUANNOTATE*, we evaluated the relationship between target delineation compliance with the published international guidelines^7^ and treatment outcomes of NPC patients undergoing radical RT.

## Methods and Materials

### Dataset

The dataset used for this study consists of anonymized computed tomography (CT) simulation scans, RT structures, and clinical data of 356 pathologically confirmed NPC patients who underwent IMRT at the Princess Margaret Cancer Center, University Health Network (UHN), Toronto, Canada, between July 2005 and August 2017. All of these patients were included in our Anthology of Outcomes^13^ and had been subject to the standard internal QA processes used for all patients treated at our center.^14^ Among these patients, 332 had a follow-up of 2 years or longer and formed the cohort for the current study. (Figure 1). Patients’ CT images contained a median of 181 slices, with each slice consisting of 512 × 512 pixels. The median slice thickness was 2.0 mm (range 2.0-3.0 mm).^15^ This study conformed to the ethical guidelines of the 1975 Declaration of Helsinki and was approved by the Health Canada and Public Health Agency of Canada (PHAC) Research Ethics Board (REB, approval #17-5871).

**Figure 1.**
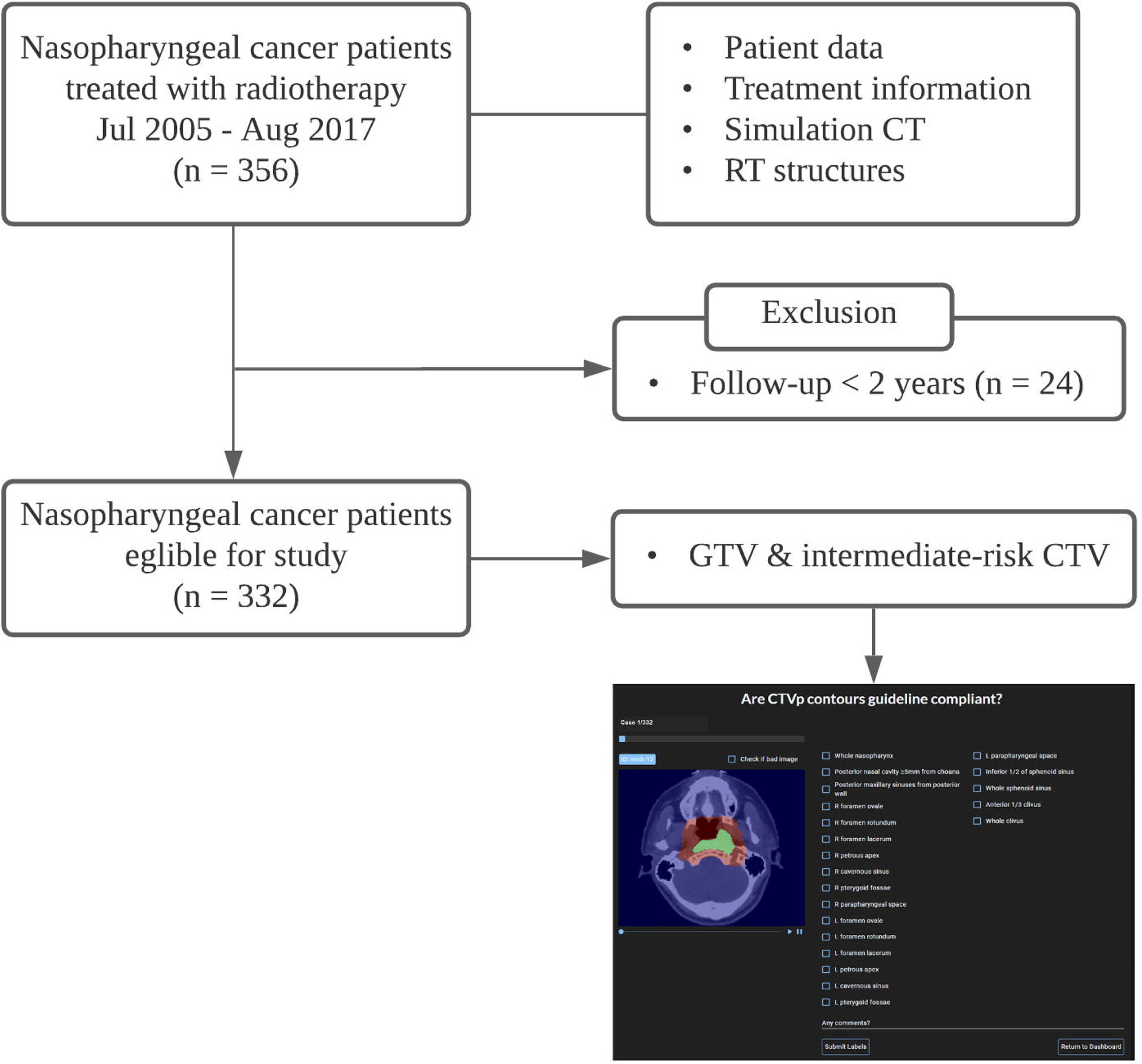
Study inclusion workflow

The contours of GTV and IR-CTV were extracted from existing RT structure sets from each patient, and a python-based framework was used to develop the web-application (design presented in Supplementary Figure 1; details about implementation provided in Supplementary Methods).

### IR-CTVp delineation compliance assessment

Two CTVs for the primary tumor (CTVp) are routinely used for NPC patients treated at Princess Margaret Cancer Center: HR-CTVp was designated as CTV70 and IR-CTVp as CTV56, and these targets correspond to CTVp1 and CTVp2 in the international guidelines for the delineation of the CTV for NPC,^7^ respectively. We loaded GTVp and IR-CTVp to *QUANNOTATE* and used binary assessments of whether or not 18 anatomic sites were included within IR-CTVp: whole nasopharynx; posterior nasal cavity ^≥^5 mm from choana; posterior maxillary sinuses from the posterior wall; right and left foramen ovale, foramen rotundum, foramen lacerum, petrous apices, pterygoid fossae, and parapharyngeal spaces; ipsilateral cavernous sinus (if T3-4), inferior 1/2 (if T1-2) or whole (if T3-4) sphenoid sinus; and anterior 1/3 (if no invasion) or whole clivus (if invasion). Compliance score was defined as the number of anatomic sites included within IR-CTVp, and a case with complete inclusion of these anatomic sites would receive a score of 18.

### Interpretation of international guidelines

Controversial issues may arise during the implementation of international guidelines in defining target volumes. For example, the expert group showed low (65%) consensus on whether or not to shave out air cavity within the HR-/IR-CTVp volumes.^7^ The guidelines recommend coverage of the ipsilateral cavernous sinus for T3-4 tumors, yet some advanced tumors may not have clear invasion of the cavernous sinus or definite lateralization. Coverage of petrous tip was recommended for all stages, yet the extent of adequate coverage for petrous apex is not clear. We defined additional criteria to help determine guideline compliance: shaving of air cavity was not allowed for sphenoid sinus but for nasal cavity and maxillary sinuses; both cavernous sinuses should be covered in case invasion or laterality is unclear for T3-4 tumors, unless T4-category is solely related to inferior extension such as to the hypopharynx; and at least anterior 1/2 of the anterior petrous apex bounded by internal auditory canal should be covered in all stages (Table 1). Blinded to the treatment outcomes, a radiation oncologist (JK) with 9 years of post-training experience, determined inclusion of the anatomic sites within IR-CTVp using *QUANNOTATE*.

**Table 1.** Criteria for intermediate risk CTVp compliance with international guidelines

| Anatomical landmarks | International guidelines | Additional criteria for guideline compliance |
| --- | --- | --- |
| Margin from GTV | GTVp + 10 mm + whole NP | None |
| Nasal cavity – Posterior part | At least 5 mm from choana | Uppermost cavity may be spared; shaving of air cavity is allowed |
| Maxillary sinuses – Posterior part | At least 5 mm from posterior wall | Uppermost sinus may be spared; shaving of air cavity is allowed |
| Posterior ethmoid sinus | Include vomer | None |
| Skull base | Cover foramina ovale, rotundum, lacerum & petrous tip | Foramina: full coverage<br>Petrous tip: at least anterior 1/2 of the anterior petrous apex bounded by internal auditory canal |
| Cavernous sinus | If T3-4 (involved side only) | Uppermost sinus may be spared; at least 5-mm margin superiorly from GTVp must be observed except for tumor abutting chiasm<br>No definite laterality: both cavernous sinuses |
| Pterygoid fossae | + | None |
| Parapharyngeal spaces | Full coverage | None |
| Sphenoid sinus | Inferior 1/2 if T1-2; whole if T3-4 | Air cavity should be included but the uppermost sinus may be spared for T3-4 |
| Clivus | 1/3 if no invasion; whole if invasion | Upper- and lowermost part may be spared, but at least 5-mm margin from invasion front must be observed except for tumor abutting brainstem |
| Minimal margin if tumor in close proximity to critical OARs | CTVp + 2 mm | None |

*Abbreviations:* GTVp = gross tumor volume of the primary tumor; CTVp = clinical target volume of the primary tumor; NP = nasopharynx; OARs = organs at risk

### Statistical analysis

The Kaplan-Meier method was used to estimate overall survival (OS) and time to local failure (TTLF) and the log-rank test was used to provide a statistical comparison of two groups. Survival was calculated from the start of RT to death due to any cause or last patient follow-up and TTLF from the start of RT to a pathologically confirmed local failure or clear radiographic progression in cases where biopsy was not indicated. Factors with p value less than 0.10 in univariable analysis (UVA) were evaluated for independence with the Cox proportional hazards model. We analyzed treatment outcomes of the whole cohort, whereas a subgroup T3-4 cases were analyzed when guidelines provided separate recommendations for locally advanced diseases. Data analysis was performed with IBM SPSS Statistics version 25.0 (BIM Corp., Armonk, NY, USA).

## Results

### Patient characteristics

Clinical characteristics of 332 NPC patients are summarized in Table 2. The median age was 52 years. Non-keratinizing undifferentiated carcinoma (Type 3) was the predominant (80.1%) histology. T3-4 tumors were diagnosed in 59.3% and cervical lymph node metastasis in 87.3% of the patients. Concurrent chemoradiation was the predominant treatment modality (87.7%) with a median total RT dose of 70.0 Gy (Table 2).

**Table 2.** Patient characteristics

| Variable |  | Number of patients | % |
| --- | --- | --- | --- |
| Age |  | Median 52 years (range, 16~88) |  |
| Gender | Male | 238 | 71.7 |
|  | Female | 94 | 28.3 |
| Performance | ECOG 0 | 236 | 71.1 |
|  | ECOG 1 | 89 | 26.8 |
|  | ECOG 2 | 3 | 0.9 |
|  | ECOG 3 | 2 | 0.6 |
|  | Unknown | 2 | 0.6 |
| Smoking history | Current | 55 | 16.6 |
|  | Ex-smoker | 79 | 23.8 |
|  | Nonsmoker | 189 | 56.9 |
|  | Unknown | 9 | 2.7 |
| Drinking history | Ex-drinker | 14 | 4.2 |
|  | Heavy (>20 drink/wk) | 11 | 3.3 |
|  | Moderate (10-20 drink/wk) | 8 | 2.4 |
|  | Light (<10 drink/wk) | 53 | 16.0 |
|  | Nondrinker | 230 | 69.3 |
|  | Unknown | 16 | 4.8 |
| Epstein-Barr virus | Positive | 241 | 72.6 |
|  | Negative | 20 | 6.0 |
|  | No test | 71 | 21.4 |
| Pathology | Type I (WHO I) | 10 | 3.0 |
|  | Type 2 (WHO IIA) | 54 | 16.3 |
|  | Type 3 (WHO IIB) | 266 | 80.1 |
|  | Neuroendocrine (Small cell) | 2 | 0.6 |
| T stage | T1 | 98 | 29.5 |
|  | T2 | 37 | 11.1 |
|  | T3 | 105 | 31.6 |
|  | T4 | 92 | 27.7 |
| N stage | N0 | 42 | 12.7 |
|  | N1 | 79 | 23.8 |
|  | N2 | 164 | 49.4 |
|  | N3 | 47 | 14.1 |
| Overall stage | I | 14 | 4.2 |
|  | II | 39 | 11.7 |
|  | III | 151 | 45.5 |
|  | IVA | 81 | 24.4 |
|  | IVB | 47 | 14.2 |
| Treatment | CCRT | 291 | 87.7 |
|  | RT alone | 41 | 12.3 |
| Total RT dose |  | Median 70.0 Gy (range, 59.4~70.0) |  |
| RT fractions |  | Median 35 fractions (range, 25~35) |  |

*Abbreviations:* ECOG = Eastern Cooperative Oncology Group; WHO = World Health Organization; CCRT = concurrent chemoradiation; RT = radiotherapy; wk = week

### Compliance of IR-CTVp to international guidelines

For all patients (n = 332), the number of cases with each anatomic site missed by IR-CTVp are as follows: 1 (0.3%) for whole nasopharynx, 2 (0.6%) for posterior nasal cavity, 2 (0.6%) for posterior maxillary sinuses, 6 (1.8%) and 4 (1.2%) for right and left foramen ovale, 14 (4.2%) and 12 (3.6%) for right and left foramen rotundum, 1 (0.3%) and 3 (0.9%) for right and left foramen lacerum, 0 and 1 (0.3%) for right and left pterygoid fossa, 0 and 1 (0.3%) for right and left parapharyngeal space, and 14 (4.2%) for the clivus. Right and left apices of the petrous bone were inadequately contoured in 32 (9.2%) and 37 (11.1%) cases, respectively, while the majority of inadequate coverage was observed in T1-2 cases. The increased number of non-contoured petrous apices in early tumors is likely due to sparing of the inner ear. Inadequate contouring of sphenoid sinus was observed in 59 (29.9%) of the T3-4 cases and 10 (7.4%) of the T1-2 cases. Contouring of cavernous sinus was recommended only for T3-4 tumors, and 38 of the 197 cases (19.3%) were inadequately covered (Figure 2A).

**Figure 2.**
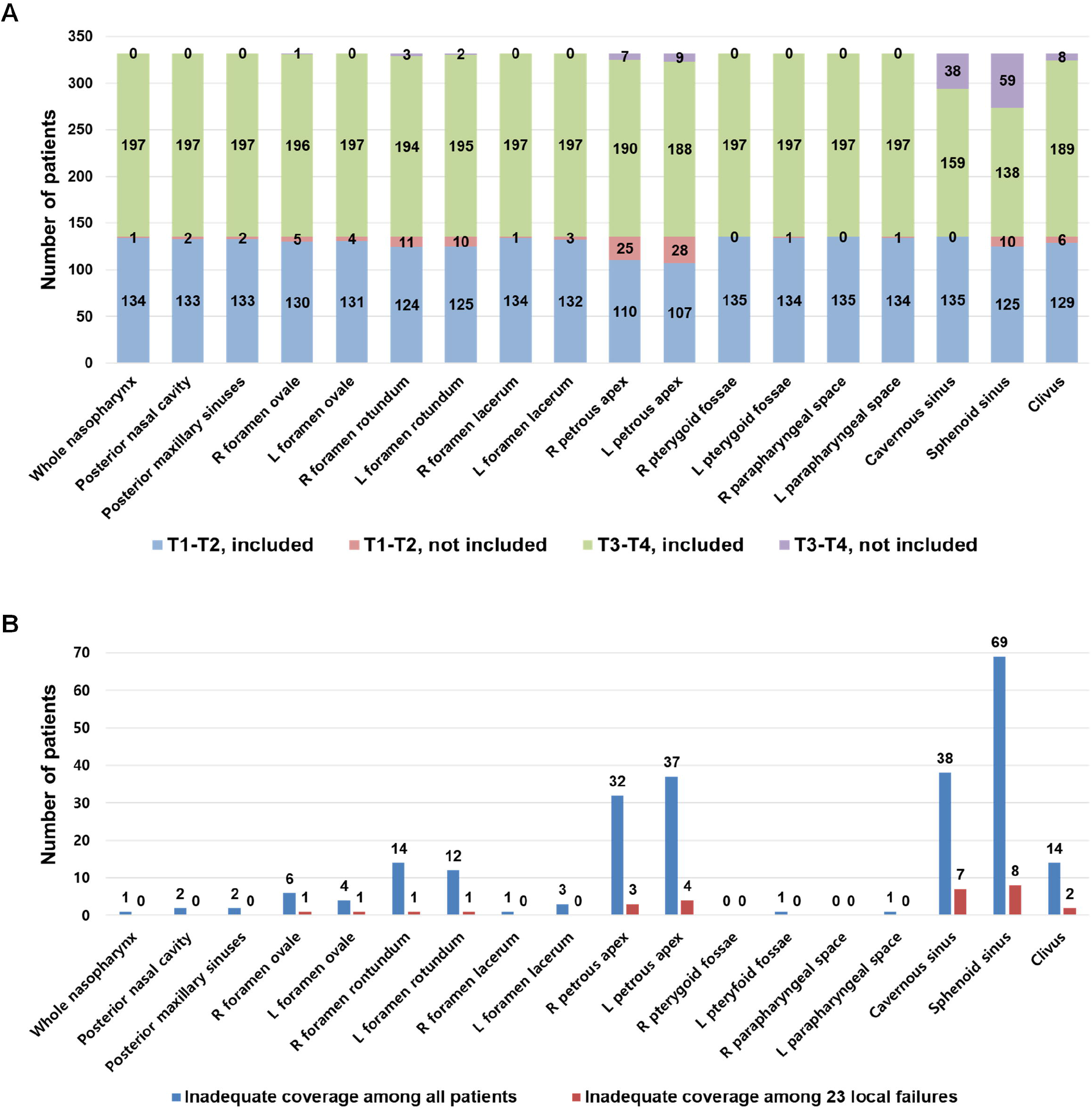
Coverage of anatomic sites by intermediate risk clinical target volume. (A) Inadequate coverage by T1-2 vs. T3-4; (B) Inadequate coverage among all patients vs. 23 local failures

### Patterns of failure

At a median follow-up of 5.6 years (range, 0.1-13.8), treatment resulted in 13 (3.9 %) local, 3 (0.9 %) regional, 37 distant (11.1 %), 7 (2.1 %) locoregional, 2 (0.6%) local and distant, 9 (2.7 %) regional and distant, and 1 (0.3%) locoregional and distant failures (Supplementary Figure 2). The ratio of non-guideline contoured anatomic sites among 23 local failures to those among all patients was highest for left (1/4) and right (1/6) foramen ovale, followed by cavernous sinus (7/38), clivus (2/14), sphenoid sinus (8/69), left (4/37) and right (3/32) petrous apices, and left (1/12) and right (1/14) foramen rotundum (Figure 2B). The compliance score (number of anatomic sites included within IR-CTVp) and number of local failures for each of the scores are shown in Supplementary Figure 3. Score of 16 or higher was achieved in 93.9% of the cases, and local failure rates increased from 5.4% for a score of 18 to 20.0% for a score of 15.

### Survival analysis

Clinical and target-compliance variables were correlated with TTLF (Table 3). T4 disease (p = 0.006) and under-contouring of the cavernous sinus (p = 0.013) showed a significant correlation with decreased TTLF and treatment with RT alone (p = 0.084) showed a marginal correlation in UVA, while these variables all proved to be independent prognostic factors in multivariable analysis (MVA). A similar trend was observed in a subgroup of T3-4 cases (n = 197), albeit cavernous sinus under-contouring (p = 0.072) was less prognostic for TTLF than T4 (p = 0.008) and RT alone (p = 0.049) in MVA (Supplementary Table 1). Nodal staging (N0-2 versus N3) was included in the analyses of recurrence-free survival (RFS) and OS, and the results of UVA and MVA are summarized in Supplementary Tables 2 and 3, respectively. T4 (p = 0.002) and N3 (p <0.0001) were independently prognostic for RFS, whereas age >53 (p = 0.023), ECOG ^≥^1 (p = 0.003), WHO I/IIa (p = 0.048), T4 (p = 0.001), N3 (p <0.0001), and RT alone (p = 0.003) showed statistically significant correlation with poor OS in MVA. Five-year TTLF and OS rates were 93.1% and 85.9% for all patients and 91.5% and 80.8% for the T3-T4 subgroup (Supplementary Figure 4). For cases conforming versus non-conforming to the guidelines, 5-year TTLF showed a significant survival difference (93.6% versus 89.1%, p = 0.013) while OS did not (87.8% versus 74.0%, p = 0.099; Figure 3).

**Table 3.** Prognostic factors for time to local failure (n = 322)

| Variable |  | No | Univariate analysis |  |  | Multivariate analysis |  |  |
| --- | --- | --- | --- | --- | --- | --- | --- | --- |
|  |  |  | HR | 95% CI | p | HR | 95% CI | p |
| Age | >53 | 157 | 2.00 | 0.87-4.64 | 0.10 |  |  |  |
|  | ≤53 | 175 | 1 |  |  |  |  |  |
| Gender | Female | 94 | 1.59 | 0.69-3.67 | 0.290 |  |  |  |
|  | Male | 238 | 1 |  |  |  |  |  |
| Performance | ECOG ≥1 | 96 | 1.61 | 0.68-3.81 | 0.28 |  |  |  |
|  | ECOG 0 | 236 | 1 |  |  |  |  |  |
| Smoking Hx | Yes/unknown | 143 | 0.78 | 0.33-1.84 | 0.57 |  |  |  |
|  | None | 189 | 1 |  |  |  |  |  |
| Alcohol Hx | Yes/unknown | 102 | 1.1 | 0.43-2.81 | 0.84 |  |  |  |
|  | None | 230 | 1 |  |  |  |  |  |
| EBV | Negative | 91 | 0.81 | 0.31-2.10 | 0.669 |  |  |  |
|  | Positive | 241 | 1 |  |  |  |  |  |
| Pathology | WHO I/IIa | 66 | 0.96 | 0.33-2.78 | 0.937 |  |  |  |
|  | WHO IIb | 266 | 1 |  |  |  |  |  |
| T stage | T4 | 92 | 3.15 | 1.39-7.14 | 0.006 | 4.02 | 1.63-9.94 | 0.003 |
|  | ≤T3 | 240 | 1 |  |  | 1 |  |  |
| Treatment | RT alone | 41 | 2.4 | 0.89-6.48 | 0.084 | 4.50 | 1.50-13.5 | 0.007 |
|  | CCRT | 291 | 1 |  |  | 1 |  |  |
| Compliance | <18 | 127 | 1.91 | 0.84-4.35 | 0.13 |  |  |  |
|  | 18 | 205 | 1 |  |  |  |  |  |
| Petrous apex | Nonconforming | 48 | 1.51 | 0.56-4.1 | 0.41 |  |  |  |
|  | Conforming | 284 | 1 |  |  |  |  |  |
| Cavernous sinus | Nonconforming | 38 | 3.07 | 1.26-7.48 | 0.013 | 2.92 | 1.19-7.19 | 0.020 |
|  | Conforming | 294 | 1 |  |  | 1 |  |  |
| Sphenoid sinus | Nonconforming | 69 | 2.04 | 0.87-4.82 | 0.10 |  |  |  |
|  | Conforming | 263 | 1 |  |  |  |  |  |

*Abbreviations:* HR = hazard ratio; CI = confidence interval; ECOG = Eastern Cooperative Oncology Group; EBV = Epstein-Barr virus; WHO = World Health Organization; RT = radiotherapy; CCRT = concurrent chemoradiation

**Figure 3.**
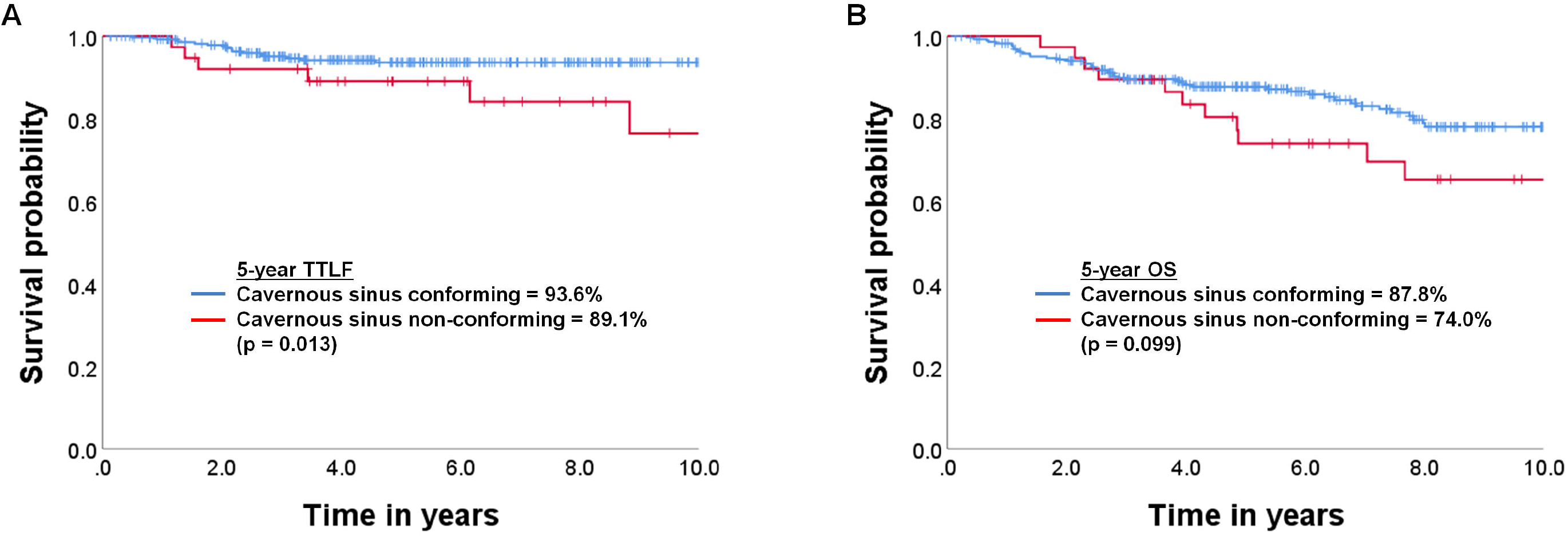
Kaplan-Meier curves for (A) time to local failure and (B) overall survival by cavernous sinus conformity

### Research reproducibility

The web-based application, QUANNOTAE, is available at https://www.quannotate.com, and all preprocessing scripts used to build the application are available at https://github.com/bhklab/quannotate.

## Discussion

Multi-institutional clinical trials involving modern RT techniques are unique in that treatment quality can be assessed by reviewing digitized medical images and associated RT structures.^16,17^ Nonetheless, QA of large-scale trials is always challenging because of time-consuming processes for collecting and reviewing hundreds or thousands of individual cases. We developed *QUANNOTATE*, a web-based QA tool for a rapid review of hundreds of NPC cases (and other primary sites). *QUANNOTATE* provides a QA platform for uploading CT simulation images and RT structures and allows reviewing target volumes with respect to published guidelines.

Advances of RT techniques in the IMRT era, particularly image guidance, facilitate more conformal (steeper dose gradients) treatments with better sparing of nearby, adjacent organs at risk (less toxicity). Ng et al. showed that when the volume within GTVp below 66.5 Gy (95% prescribed dose) was greater than the cutoff volume of 3.4 cm^3^, the 5-year local failure-free survival decreased from 90% to 54%.^18^ This suggests that survival, as function of target coverage, is highly non-linear, where even a small “cold spot” (i.e., 3.4 cm^3^ implies sphere of radius 0.93 cm) can have an enormous impact (i.e., 40% reduction in survival from 90 to 54%).

With a median follow-up of 5.6 years in the current study, the local recurrence rate of 6.9% for NPC is comparable with previously published IMRT data.^19,20^ Nevertheless, inadequate coverage of the cavernous sinus was observed in 19.3% of T3-4 cases and was an independent prognostic factor for TTLF. The importance of including the cavernous sinus within IR-CTVp was not universally recognized: Among the 6 studies reviewed by the international guidelines committee,^7^ coverage of the cavernous sinus was not stated in 2 studies, RTOG 0225^2^ and the Chinese guidelines.^21^ The recommendation to include the involved-side cavernous sinus for T3-4 tumor was based on the findings by Liang et al. where the cumulative incidence rate of cavernous sinus invasion was 17.4%; however, the structure was only at high risk when the tumor infiltrated the petrous apex or the foramen lacerum.^22^ Liang et al also demonstrated the importance of multiple infiltration routes to cavernous sinus: the rate of cavernous sinus invasion through a single route was 39.4% (foramen ovale being the most common) compared with 60.7% when two or more routes (foramen ovale and lacerum being the most common) were involved. Our data support the international guidelines’ recommendations on the coverage of the cavernous sinus. High local failure rates associated with inadequate coverage of other anatomic sites including foramen ovale and sphenoid sinus suggest that, with a larger study cohort, we may be able to “rank” these anatomic sties in order of importance in delineating target volumes.

Medical imaging provides the ability to detect and localize regions of interests (ROIs) and important changes to treatment, hence imaging biomarker studies are rapidly gaining popularity. Radiomics studies extract features from a large number of radiological medical images using diverse computational approaches.^23-25^ While measures are taken to ensure reproducibility of the results via standardizing processes of image acquisition and data analysis,^26,27^ the impact of treatment quality is often overlooked in radiomics studies, potentially confounding the search for robust biomarkers. With respect to optimizing plans, standardizing treatment to follow known, accepted international guidelines minimize variance. These changes are “low hanging fruit” and, arguably, the simplest, most impactful change one can do to improve treatment outcomes. If a chemotherapy agent demonstrated a comparable positive effect as rigorous treatment QA, it would have been adopted without question.

Our results suggest that radiomics analysis of published datasets should be preceded by detailed QA analysis to ensure outcomes are not confounded due to variance in treatment-related factors as opposed to tumor factors. This tool could also be used as a measure for treatment quality but in a more general sense. For example, the proportion of plans that conform within the guidelines could be used as quality metric at the level of the individual, site group, department, or even institution.

The potential use of *QUANNOTATE* in central review of RT targets/plans in clinical trials and patterns-of-radiotherapy-practice studies has been previously mentioned. The presentation format of *QUANNOTATE* can be modified according to the objectives of its application, and we can suggest several areas where use of *QUANNOTATE* can be beneficial. First, the target delineation QA platform can readily be utilized for assessment of anatomical understanding and pathway of tumor spread and can be an effective tool for teaching and assessing medical students and radiation oncology residents. Second, RT planning studies often review and correlate radiation dose distribution with treatment outcomes. Dose distribution data stored in RT planning station can readily be transported onto *QUANNOTATE* and reviewed with corresponding RT target volumes. Third, upgraded with additional viewing windows and a toggle switch for image selection, *QUANNOTATE* can be used to compare ROIs between serially taken images and this can be an effective tool for evaluating treatment responses in studies with large sample sizes. Our goal is to implement artificial intelligence-based image recognition in the *QUANNOTATE* platform, and this will open doors to many more applications.

This study is subject to the limitations of all retrospective and single-institution studies, such as selection bias, small number of events, and observer bias. The objective of this study was to introduce an online method to review large image datasets in an easily accessible format. Among its potential applications, we first used it as a QA tool for target delineation in NPC patients. We focused on the compliance of IR-CTVp with the international guidelines, thus the outcome of interest was mainly local failure. With high compliance of target volumes and a small number of events (23 local failures), we lacked power to examine the significance of other anatomic sites. In accordance with the study by Liang et al., one of 4∼6 (17∼25%) cases with inadequate coverage of foramen ovale resulted in local failures in the current study. A validation study with a large external cohort would increase the power to examine the impact of undercoverage of other anatomic sites. Secondly, compliance of IR-CTVp with the international guidelines was reviewed by only one experienced radiation oncologist. We implemented additional criteria of compliance to minimize observer bias by the reviewer. A validation study with an external cohort and inter-observer variation analysis is underway.

## Conclusions

We developed a web-based QA tool for a contour review of target volumes for a large number of NPC patients. Despite a high compliance with the international guidelines and internal QA processes, *QUANNOTATE* identified inadequate coverage of the cavernous sinus was correlated with local failure.

## Data Availability

https://www.quannotate.com

## Figure legends

Supplementary Figure 1. (A) Simple software diagram of the *QUANNOTATE* application. (B) Web-based review of RT target delineation using *QUANNOTATE*.

Supplementary Figure 2. Patterns of failure

Supplementary Figure 3. Compliance scores and corresponding local failure rates

Supplementary Figure 4. Kaplan-Meier curves for (A) time to local failure for all patients (B) overall survival for all patients, (C) time to local failure for T3-T4, and (D) overall survival for T3-T4

