## Supplementary Information for "Development of web-based quality-assurance tool for radiotherapy target delineation for head and neck cancer: quality evaluation of nasopharyngeal carcinoma"

**Supplementary Figures**

Supplementary Figure 1A. Simple software diagram of the *QUANNOTATE* application; 1B Web-based review of RT target delineation using *QUANNOTATE*

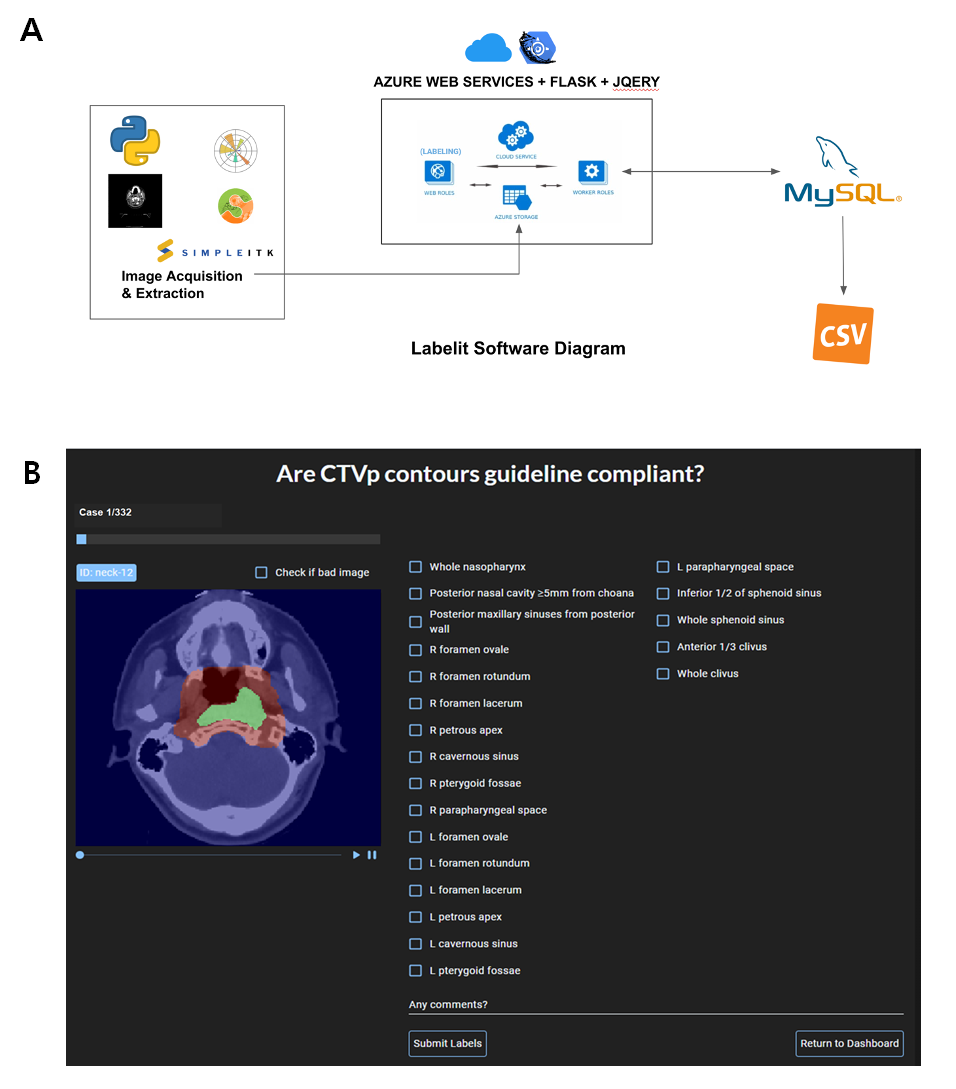

Supplementary Figure 2. Patterns of failure

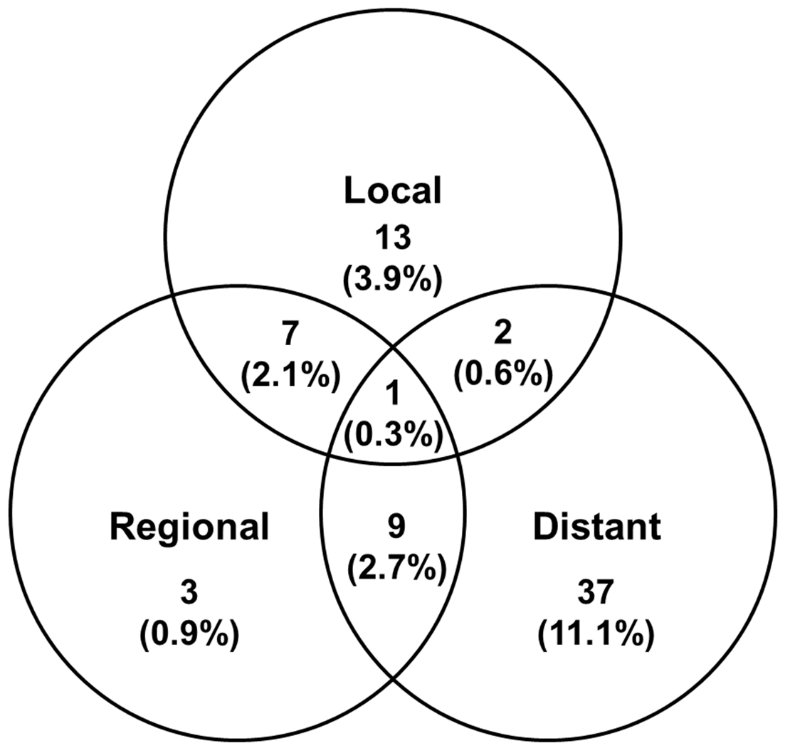

Supplementary Figure 3. Compliance scores and corresponding local failure rates

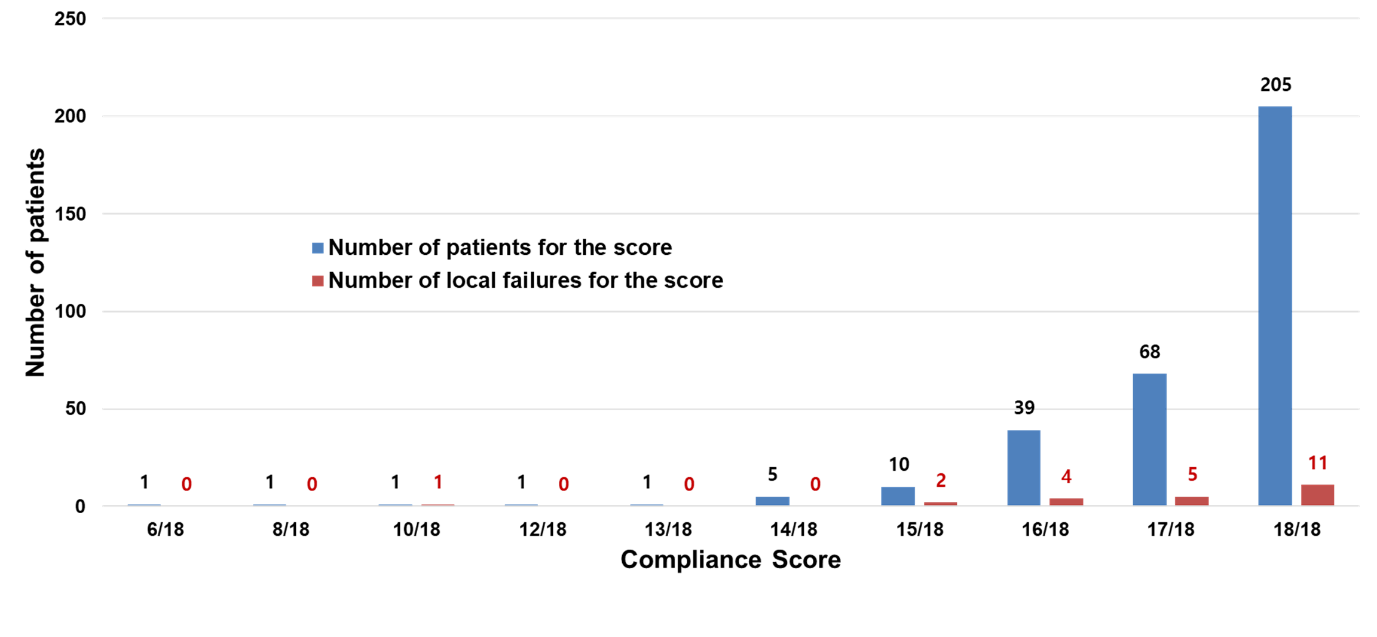

Supplementary Figure 4. Kaplan-Meier curves for (A) time to local failure for all patients (B) overall survival for all patients, (C) time to local failure for T3-T4, and (D) overall survival for T3-T4
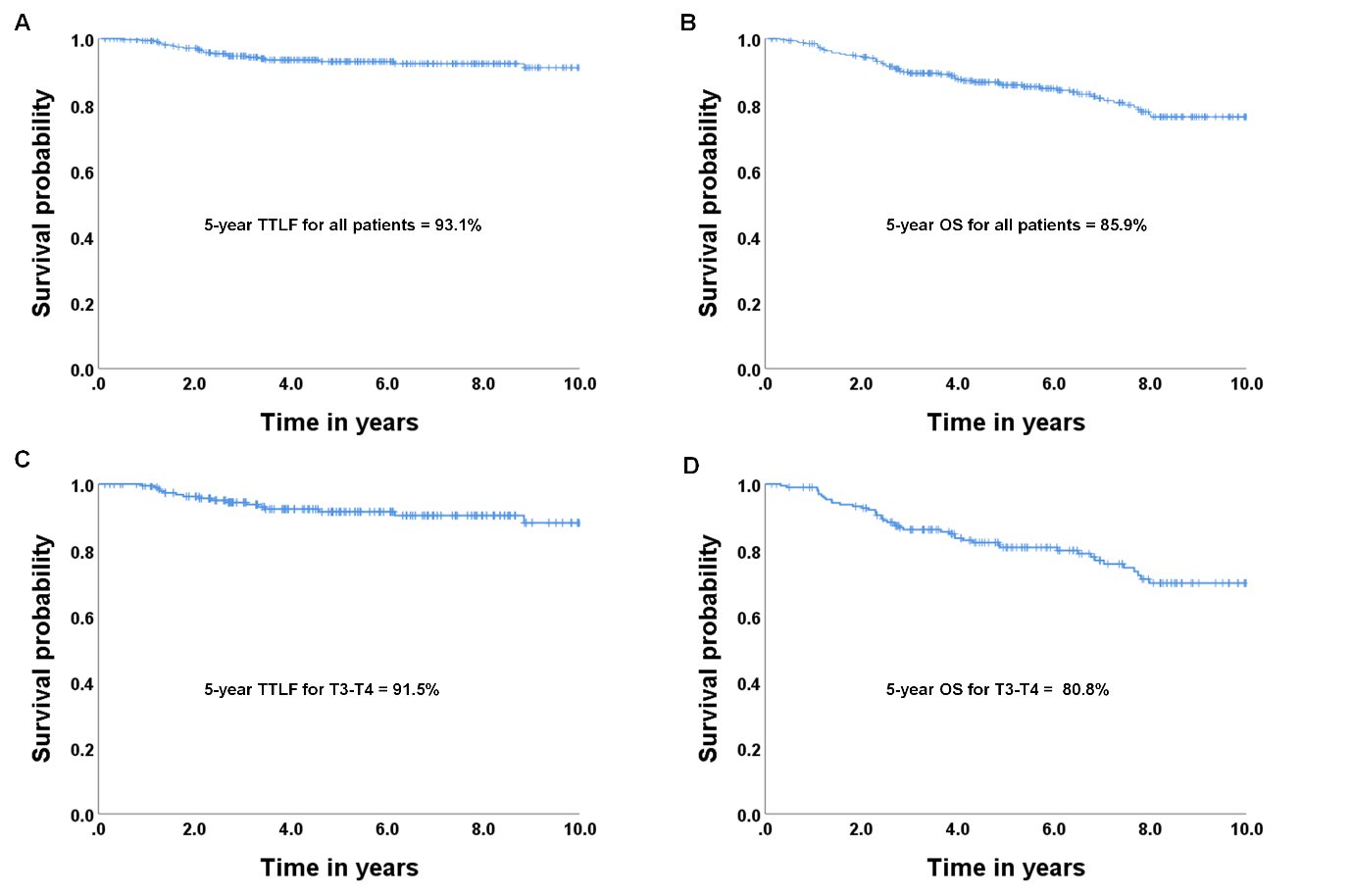

**Supplementary Tables**

Supplementary Table 1. Prognostic factors for time to local failure among T3-4 cases (n = 197)

| Variable |  | No | Univariate analysis | | |  | Multivariate analysis | | |
| --- | --- | --- | --- | --- | --- | --- | --- | --- | --- |
|  |  |  | HR | 95% CI | p |  | HR | 95% CI | p |
| Age | >53 | 91 | 2.50 | 0.92-6.76 | 0.072 |  | 1.02 | 0.97-1.07 | 0.47 |
|  | ≤53 | 106 | 1 |  |  |  | 1 |  |  |
| Gender | Female | 57 | 1.19 | 0.44-3.22 | 0.740 |  |  |  |  |
|  | Male | 140 | 1 |  |  |  |  |  |  |
| Performance | ECOG ≥1 | 66 | 1.0 | 0.35-2.85 | 1.00 |  |  |  |  |
|  | ECOG 0 | 131 | 1 |  |  |  |  |  |  |
| Smoking Hx | Yes | 83 | 0.79 | 0.29-2.14 | 0.64 |  |  |  |  |
|  | None | 114 | 1 |  |  |  |  |  |  |
| Alcohol Hx | Yes | 66 | 1.38 | 0.51-3.75 | 0.64 |  |  |  |  |
|  | None | 131 | 1 |  |  |  |  |  |  |
| EBV | Negative | 52 | 0.73 | 0.23-2.28 | 0.585 |  |  |  |  |
|  | Positive | 145 | 1 |  |  |  |  |  |  |
| Pathology | WHO I/IIa | 42 | 1.24 | 0.38-4.08 | 0.720 |  |  |  |  |
|  | WHO IIb | 155 | 1 |  |  |  |  |  |  |
| T stage | T4 | 92 | 2.95 | 1.04-8.39 | 0.042 |  | 4.68 | 1.5-14.57 | 0.008 |
|  | ≤T3 | 105 | 1 |  |  |  | 1 |  |  |
| Treatment | RT alone | 15 | 4.83 | 1.34-17.4 | 0.016 |  | 5.51 | 1.01-30.1 | 0.049 |
|  | CCRT | 182 | 1 |  |  |  | 1 |  |  |
| Compliance | <18 | 86 | 1.62 | 0.62-4.26 | 0.33 |  |  |  |  |
|  | 18 | 111 | 1 |  |  |  |  |  |  |
| Petrous apex | Nonconforming | 15 | 1.55 | 0.35-6.81 | 0.56 |  |  |  |  |
|  | Conforming | 182 | 1 |  |  |  |  |  |  |
| Cavernous sinus | Nonconforming | 38 | 2.46 | 0.93-6.49 | 0.069 |  | 2.52 | 0.92-6.89 | 0.072 |
|  | Conforming | 159 | 1 |  |  |  | 1 |  |  |
| Sphenoid sinus | Nonconforming | 59 | 1.57 | 0.6-4.13 | 0.36 |  |  |  |  |
|  | Conforming | 138 | 1 |  |  |  |  |  |  |

*Abbreviations:* HR = hazard ratio; CI = confidence interval; ECOG = Eastern Cooperative Oncology Group; EBV = Epstein-Barr virus; WHO = World Health Organization; RT = radiotherapy; CCRT = concurrent chemoradiotherapy

Supplementary Table 2. Prognostic factors for recurrence-free survival (n = 322)

| Variable |  | No | Univariate analysis | | |  | Multivariate analysis | | |
| --- | --- | --- | --- | --- | --- | --- | --- | --- | --- |
|  |  |  | HR | 95% CI | p |  | HR | 95% CI | p |
| Age | >53 | 157 | 1.34 | 0.84-2.12 | 0.220 |  |  |  |  |
|  | ≤53 | 175 | 1 |  |  |  |  |  |  |
| Gender | Female | 94 | 0.83 | 0.44-1.32 | 0.480 |  |  |  |  |
|  | Male | 238 | 1 |  |  |  |  |  |  |
| Performance | ECOG ≥1 | 96 | 1.46 | 0.89-2.40 | 0.130 |  |  |  |  |
|  | ECOG 0 | 236 | 1 |  |  |  |  |  |  |
| Smoking Hx | Yes | 143 | 0.91 | 0.57-1.47 | 0.710 |  |  |  |  |
|  | None | 189 | 1 |  |  |  |  |  |  |
| Alcohol Hx | Yes | 102 | 0.98 | 0.57-1.69 | 0.940 |  |  |  |  |
|  | None | 230 | 1 |  |  |  |  |  |  |
| EBV | Negative | 91 | 1.13 | 0.68-1.87 | 0.649 |  |  |  |  |
|  | Positive | 241 | 1 |  |  |  |  |  |  |
| Pathology | WHO I/IIa | 66 | 0.97 | 0.54-1.76 | 0.923 |  |  |  |  |
|  | WHO IIb | 266 | 1 |  |  |  |  |  |  |
| T stage | T4 | 92 | 2.12 | 1.34-3.35 | <0.001 |  | 2.14 | 1.33-3.43 | 0.002 |
|  | ≤T3 | 240 | 1 |  |  |  |  | 1 |  |
| N stage | N3 | 47 | 4.32 | 2.13-8.75 | <0.001 |  | 2.62 | 1.55-4.42 | <0.001 |
|  | N0-2 | 285 | 1 |  |  |  |  | 1 |  |
| Treatment | RT alone | 41 | 1.39 | 0.73-2.64 | 0.320 |  |  |  |  |
|  | CCRT | 291 | 1 |  |  |  |  |  |  |
| Compliance | <18 | 127 | 1.38 | 0.87-2.18 | 0.170 |  |  |  |  |
|  | 18 | 205 | 1 |  |  |  |  |  |  |
| Petrous apex | Nonconforming | 48 | 1.27 | 0.69-2.35 | 0.440 |  |  |  |  |
|  | Conforming | 284 | 1 |  |  |  |  |  |  |
| Cavernous sinus | Nonconforming | 38 | 1.42 | 0.8-2.54 | 0.242 |  |  |  |  |
|  | Conforming | 294 | 1 |  |  |  |  |  |  |
| Sphenoid sinus | Nonconforming | 69 | 1.84 | 1.13-3.00 | 0.014 |  | 1.56 | 0.94-2.59 | 0.085 |
|  | Conforming | 263 | 1 |  |  |  |  | 1 |  |

*Abbreviations:* HR = hazard ratio; CI = confidence interval; ECOG = Eastern Cooperative Oncology Group; EBV = Epstein-Barr virus; WHO = World Health Organization; RT = radiotherapy; CCRT = concurrent chemoradiotherapy

Supplementary Table 3. Prognostic factors for overall survival (n = 322)

| Variable |  | No | Univariate analysis | | |  | Multivariate analysis | | |
| --- | --- | --- | --- | --- | --- | --- | --- | --- | --- |
|  |  |  | HR | 95% CI | p |  | HR | 95% CI | p |
| Age | >53 | 157 | 2.29 | 1.35-3.87 | 0.002 |  | 1.92 | 1.09-3.39 | 0.023 |
|  | ≤53 | 175 | 1 |  |  |  | 1 |  |  |
| Gender | Female | 94 | 0.70 | 0.39-1.28 | 0.241 |  |  |  |  |
|  | Male | 238 | 1 |  |  |  |  |  |  |
| Performance | ECOG ≥1 | 96 | 2.46 | 1.46-4.13 | <0.001 |  | 2.23 | 1.33-3.76 | 0.003 |
|  | ECOG 0 | 236 | 1 |  |  |  | 1 |  |  |
| Smoking Hx | Yes | 143 | 1.32 | 0.79-2.21 | 0.285 |  |  |  |  |
|  | None | 189 | 1 |  |  |  |  |  |  |
| Alcohol Hx | Yes | 102 | 1.19 | 0.66-2.13 | 0.571 |  |  |  |  |
|  | None | 230 | 1 |  |  |  |  |  |  |
| EBV | Negative | 91 | 1.40 | 0.83-2.38 | 0.205 |  |  |  |  |
|  | Positive | 241 | 1 |  |  |  |  |  |  |
| Pathology | WHO I/IIa | 66 | 1.93 | 0.99-3.77 | 0.055 |  | 1.80 | 1.01-3.22 | 0.048 |
|  | WHO IIb | 266 | 1 |  |  |  | 1 |  |  |
| T stage | T4 | 92 | 1.98 | 1.19-3.31 | 0.009 |  | 2.53 | 1.44-4.43 | 0.001 |
|  | ≤T3 | 240 | 1 |  |  |  | 1 |  |  |
| N stage | N3 | 47 | 4.26 | 2.02-8.97 | <0.001 |  | 3.82 | 2.15-6.79 | <0.001 |
|  | N0-2 | 285 | 1 |  |  |  | 1 |  |  |
| Treatment | RT alone | 41 | 2.24 | 1.19-4.23 | 0.013 |  | 2.95 | 1.44-6.05 | 0.003 |
|  | CCRT | 291 | 1 |  |  |  | 1 |  |  |
| Compliance | <18 | 127 | 1.24 | 0.74-2.07 | 0.415 |  |  |  |  |
|  | 18 | 205 | 1 |  |  |  |  |  |  |
| Petrous apex | Nonconforming | 48 | 1.03 | 0.52-2.04 | 0.938 |  |  |  |  |
|  | Conforming | 284 | 1 |  |  |  |  |  |  |
| Cavernous sinus | Nonconforming | 38 | 1.70 | 0.89-3.21 | 0.099 |  | 1.26 | 0.66-2.42 | 0.488 |
|  | Conforming | 294 | 1 |  |  |  | 1 |  |  |
| Sphenoid sinus | Nonconforming | 69 | 1.50 | 0.86-2.64 | 0.160 |  |  |  |  |
|  | Conforming | 263 | 1 |  |  |  |  |  |  |

*Abbreviations:* HR = hazard ratio; CI = confidence interval; ECOG = Eastern Cooperative Oncology Group; EBV = Epstein-Barr virus; WHO = World Health Organization; RT = radiotherapy; CCRT = concurrent chemoradiotherapy

**Supplementary Methods**

*Dataset extraction*

The contours contained in the RT structure set were parsed to find IR-CTV contours for each patient. There were variants in naming for each target volume within the RT structure set. A series of regular expression strings was created to standardize extraction of the IR-CTV target of interest for each patient. Images and IR-CTV masks were extracted and de-identified using an internal image extraction protocol written in python. Images and masks were initially saved with original spacing. After initial extraction, the NPC cohort was further processed to extract all the slices from each patient into individual .png images which were saved in a corresponding patient-labeled folder. Major dependencies used include pydicom, simpleITK, dicom-contour, numpy and matplotlib. All preprocessing scripts can be accessed at https://github.com/bhklab/QUANNOTATE.

*Web-application development*

Flask was used as the python based microframework of choice for the base, proof of concept version of *QUANNOTATE*. An in-page JavaScript extension was built and used to parse through the sequential slices of each patient. Patient folders were compressed and deployed with the web-app to Azure App Service where the web-app was hosted. Images were de-compressed after deployment in the static folder of the *QUANNOTATE* app directory. A relational MySQL database was used to store labeling information for each user. Labels were then exported to .csv and used in further analysis (Supplementary Figure 1A).

*QUANNOTATE design and instructions*

We developed *QUANNOTATE* to rapidly review RT target contours for large numbers of patients. The application is composed of two sections, a window showing RT target contours on a CT slice with a scroll bar to view the whole range of CT slices, and a list of anatomic sites with checkboxes to mark inclusion within the target contour (Supplementary Figure 1B). For each case, Digital Imaging and Communications in Medicine (DICOM) images of simulation CT scan were de-identified and anonymized and loaded onto the website with corresponding RT target contours. Upon completion of reviewing a case, the list of marked anatomic sites are stored and the next case is loaded onto the viewing window.
