## Supplementary figures and images for "Development of web-based quality-assurance tool for radiotherapy target delineation for head and neck cancer: quality evaluation of nasopharyngeal carcinoma"

### Supplementary Figure 1

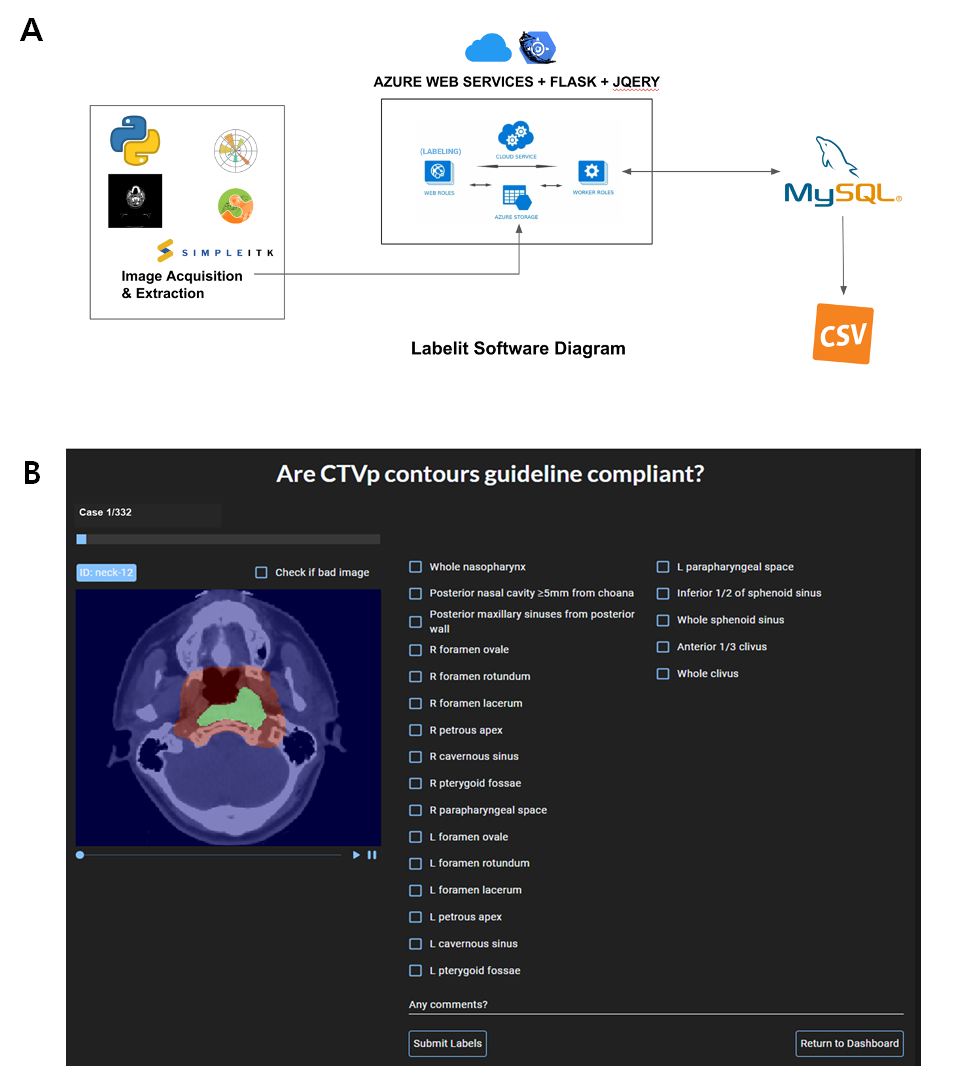

### Supplementary Figure 2

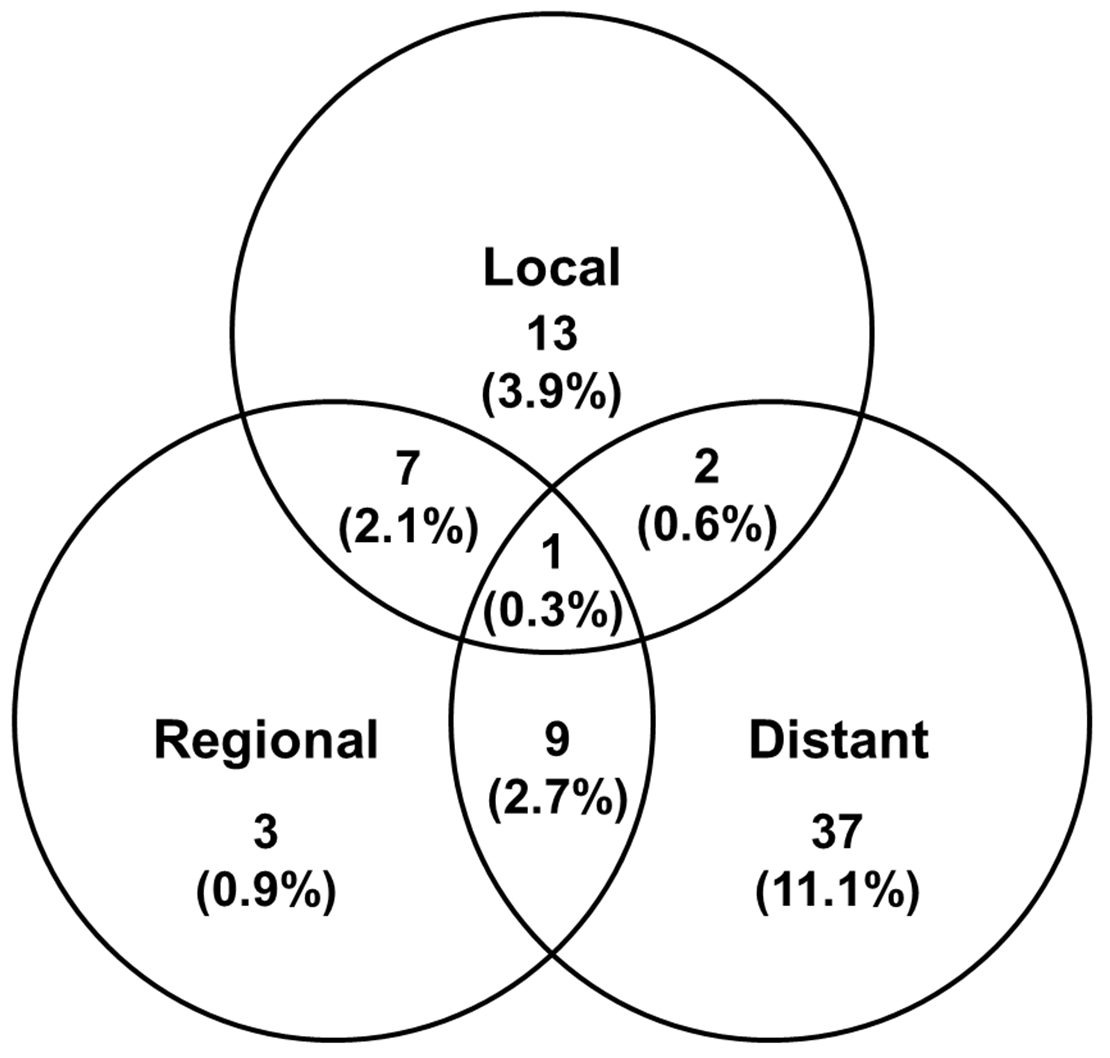

### Supplementary Figure 3

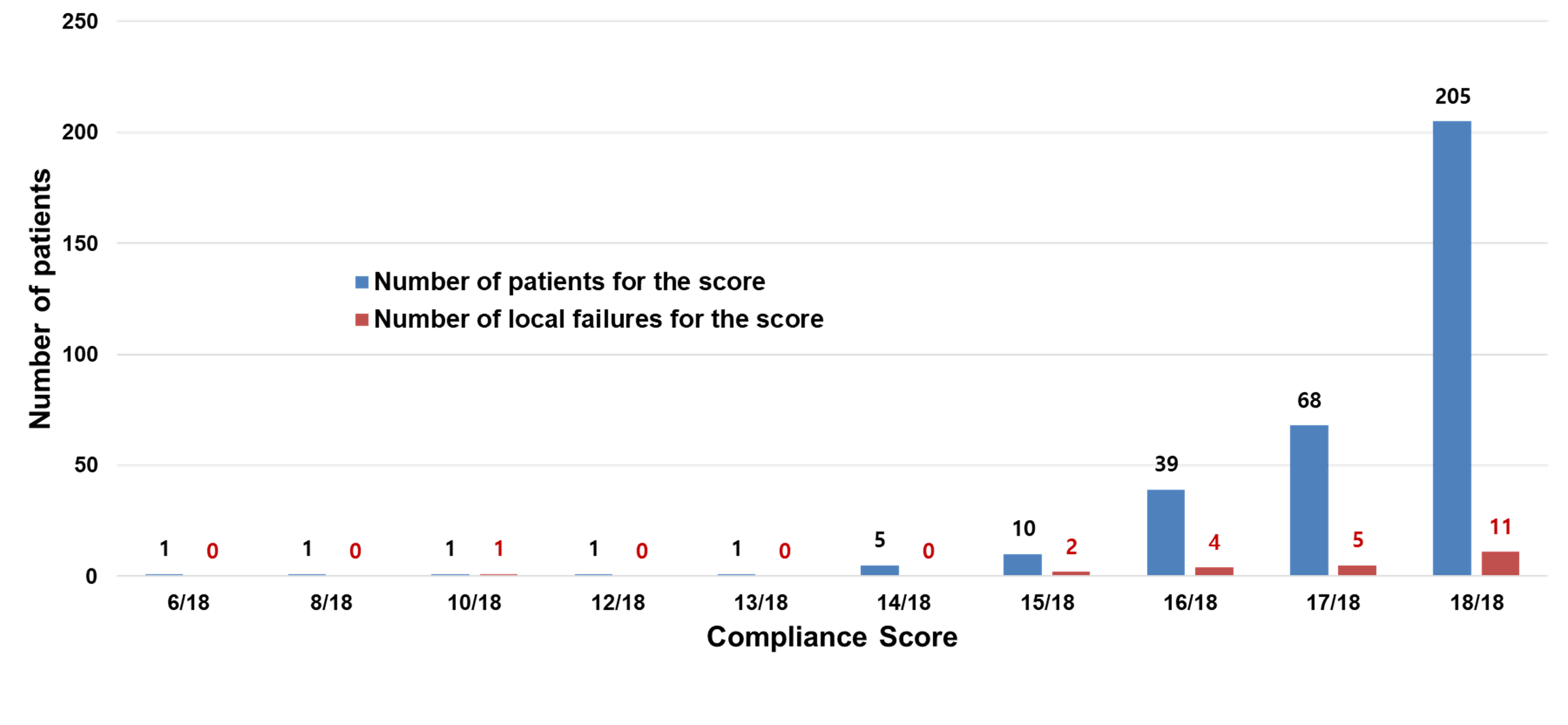

### Supplementary Figure 4

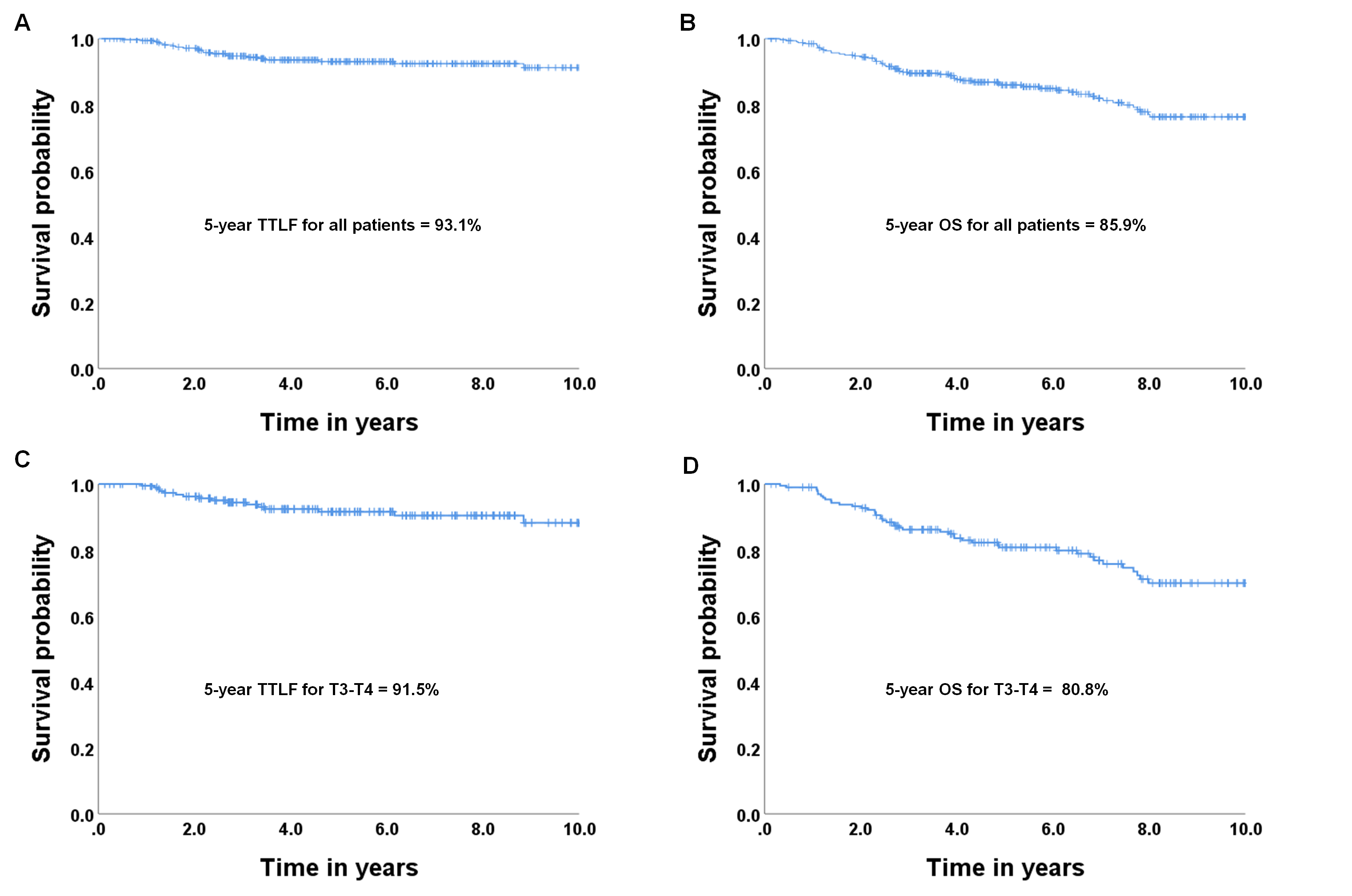
